# Special Olympics global report on COVID-19 vaccination and reasons not to vaccinate among adults with intellectual disabilities

**DOI:** 10.1101/2022.11.16.22282406

**Authors:** Andrew E. Lincoln, Alicia M. Dixon-Ibarra, John P. Hanley, Ashlyn L. Smith, Kiki Martin, Alicia Bazzano

## Abstract

**Introduction:** The COVID-19 pandemic has disproportionately affected people with intellectual disabilities worldwide. The objective of this study was to identify global rates of COVID-19 vaccination and reasons not to vaccinate among adults with intellectual disabilities (ID) associated with country economic income levels.

**Methods:** The Special Olympics COVID-19 online survey was administered in January-February 2022 to adults with ID from 138 countries. Descriptive analyses of survey responses include 95% margins of error. Logistic regression and Pearson Chi-squared tests were calculated to assess associations with predictive variables for vaccination using R 4.1.2 software.

**Results:** Participants (n=3560) represented 18 low (n=410), 35 lower-middle (n=1182), 41 upper-middle (n=837), and 44 high (n=1131) income countries. Globally, 76% (74.8-77.6%) received a COVID-19 vaccination while 49.5% (47.9-51.2%) received a COVID-19 booster. Upper-middle (93% (91.2-94.7%)) and high-income country (94% (92.1-95.0%)) participants had the highest rates of vaccination while low-income countries had the lowest rates (38% (33.3-42.7%)). In multivariate regression models, country economic income level (OR = 3.12, 95% CI [2.81, 3.48]), age (OR = 1.04, 95% CI [1.03, 1.05]), and living with family (OR = 0.70, 95% CI [0.53, 0.92]) were associated with vaccination. Among LLMICs, the major reason for not vaccinating was lack of access (41.2% (29.5-52.9%)). Globally, concerns about side effects (42%, (36.5-48.1%)) and parent/guardian not wanting the adult with ID to vaccinate (32% (26.1-37.0%)) were the most common reasons for not vaccinating.

**Conclusion:** Adults with ID from low and low-middle income countries reported fewer COVID-19 vaccinations, suggesting reduced access and availability of resources in these countries. Globally, COVID-19 vaccination levels among adults with ID were higher than the general population. Interventions should address the increased risk of infection for those in congregate living situations and family caregiver apprehension to vaccinate this high-risk population.

## Introduction

The COVID-19 pandemic has impacted people with intellectual disabilities (ID) across the world. Intellectual disability originates during early human development and is defined by significant limitations in intellectual functioning and adaptive functioning in conceptual, social, and practical domains (1). People with ID have a higher risk of contracting COVID-19, have poorer health outcomes than the general public, and are on average six times more likely than the general population to die from COVID-19 following hospital admission (2–5). COVID-19 was the leading cause of death among those with intellectual and developmental disabilities in 2020 (6). People with specific diagnoses associated with ID are particularly prone to increased risk of hospitalization and death, including those with Down syndrome, Autism, cerebral palsy, and fragile X syndrome (7–11). There are numerous reasons why people with ID are particularly vulnerable to experiencing severe COVID-19 illness and death (12). Specifically, people with ID may have difficulty accessing care as readily as the general population (13,14). Some live in congregate settings in which COVID-19 has spread more easily (15,16). People with ID are also much more likely to have preexisting conditions that raise the likelihood of experiencing severe outcomes following COVID-19 infection, such as hereditary and cardiac conditions, inborn errors of metabolism, respiratory conditions, and obesity (12). Finally, people with ID may have more challenges with risk mitigation measures, such as masking and distancing (17).

The increased risk of COVID-19 and its consequences make people with ID a particularly important group for vaccination. However, global use of vaccines during the COVID-19 pandemic has differed based on regional public health efforts, allocation of vaccines, type of vaccine, and vaccine acceptance. In low-income nations, challenges in COVID-19 testing and vaccine availability, distribution, and administration are barriers to controlling the spread of the epidemic (18). Further, COVID-19 protocols for preventing, mitigating, and treating/managing infection are largely based upon the general population without considering their effects on persons with ID. These practices have resulted in pandemic-related disparities among those with ID, including poorer physical and mental health (19). Overall, social disparities may have an outsized effect on health outcomes, healthcare access, and basic social determinants of health including food availability, safe housing, and school and work accommodations.

Enhancing equity of persons with ID in the tracking of vaccine uptake and pandemic-related health outcomes is paramount (20). This can be achieved through partnerships with the disability community and disability organizations. Special Olympics is an international non-profit organization that provides sports competition and promotes health, education and leadership opportunities to over five million people with ID in 200 countries (21). Early in the COVID-19 pandemic, Special Olympics recognized the potential increased risk for people with ID and created resources to help participants stay healthy and maintain connections among the Special Olympics community. Special Olympics compiled and disseminated accessible resources to program participants and the larger community on topics such as healthy behaviors during COVID-19 (e.g., handwashing, wearing a face mask), resources designed to help participants maintain fitness levels at home, and other virtual activities to maintain social connections with participants, coaches, and friends (22). Prevention activities and the promotion of COVID-19 vaccination were largely carried out at the local level across the world, including low- and high-income countries. Among the general population, COVID-19 infection, use of prevention strategies, and vaccination varied by country economic levels (23–25). Special Olympics’s presence across countries with varying economic levels drove the need to better understand global characteristics of the COVID-19 pandemic among adults with ID.

The objective of this study was to investigate global rates of COVID-19 vaccination and reasons not to vaccinate among adults with intellectual disabilities relative to World Bank country economic income levels.

## Methods

The Special Olympics COVID-19 survey was conducted with an online sample of adults with intellectual disabilities 18 years or older who participated in Special Olympics sport, health, schools/education, or leadership programming in 138 countries. Local staff distributed the survey to Special Olympics participants and families. The survey was developed via the Qualtrics platform (Qualtrics, Provo, UT) and remained open for responses from January through February 2022. Questions were self-reported by adults with ID, though proxy responses from family members/caregivers or staff/volunteers were also accepted. The voluntary survey included questions regarding demographics, disability characteristics, COVID-19 vaccination, and reasons for non-vaccination. COVID-19 vaccination was measured by asking participants if they received a COVID-19 vaccine. Those who had not received a COVID-19 vaccination or planned to receive it were asked what the reasons were for their decision.

Data across 138 countries were categorized by the World Bank economic income levels of low-income country (LIC), lower-middle income country (LMIC), upper-middle income country (UMIC), and high-income country (HIC) for primary analyses (26). Analyses were conducted to examine COVID-19 vaccination by diagnosis. Descriptive statistics were calculated as count and proportion of responses. For all proportions, 95% margins of error were calculated (27). Logistic regression was performed to highlight the associations between demographic variables and country economic income level with COVID-19 vaccination rates. We chose a priori to include demographic variables in the regression model while considering other interpersonal (e.g., living situation) and macro (e.g., country economic level) variables. Pearson Chi-squared tests estimated differences in vaccine acceptance across low and lower-middle income countries (LLMIC) compared to upper-middle and high-income countries (UMHIC). Analyses were performed using R version 4.1.2 (R Foundation for Statistical Computing; Vienna, Austria). This investigation was reviewed by the Oregon State University Institutional Review Board, which determined that it does not meet the definition of human subjects research under the regulations set forth by the Department of Health and Human Services 45 CFR 46.

## Results

In total, 3560 individuals from 138 countries (181 country and state-level Special Olympics Programs) responded to the survey. Survey respondents represented 18 LICs (n=410), 35 LMICs (n=1182), 41 UMICs (n=837), and 44 HICs (n=1131). Survey respondents included adults with ID (38.3%), family/caregiver responding on behalf of an adult with ID (39.1%), and program staff/volunteers responding on behalf of an adult and ID (22.6%). The median age across adults with ID was 25 years (Q1-Q3: 21-33 years) with 59.0% male. Most adults with ID (82.9%) lived with family. Participant demographics stratified by World Bank country economic levels are presented in Table 1.

**Table 1.** Global Survey Respondent Characteristics by World Bank Economic Income Level (%, 95% Margin of Error)^a^.

|  | Low-Income<br>(n=410) | Lower-Middle<br>Income<br>(n=1182) | Upper-Middle<br>Income<br>(n=837) | High-Income<br>(n=1131) | Total<br>(n=3560) |
| --- | --- | --- | --- | --- | --- |
| Median Age<br>(years), (Q1-Q3) | 23<br>(20-26) | 24<br>(20-28) | 26<br>(21-34) | 30<br>(24-38) | 25<br>(21-33) |
| Sex |  |  |  |  |  |
| Male | 57.8%<br>(53.0-62.6%) | 60.0%<br>(57.2-62.8%) | 62.0%<br>(58.7-65.3%) | 56.1%<br>(53.3-59.0%) | 59.0%<br>(57.4-60.6%) |
| Female | 42.0%<br>(37.2-46.7%) | 39.7%<br>(36.9-42.5%) | 37.5%<br>(34.2-40.8%) | 43.3%<br>(40.4-46.2%) | 40.6%<br>(39.0-42.2%) |
| Living arrangement |  |  |  |  |  |
| Live with family | 86.3%<br>(83.0-89.7%) | 86.5%<br>(84.5-88.4%) | 87.1%<br>(84.8-89.4%) | 74.8%<br>(72.3-77.3%) | 82.9%<br>(81.7-84.1%) |
| Live on own | 5.1%<br>(3.0-7.3%) | 4.7%<br>(3.5-5.9%) | 5.4%<br>(3.8-6.9%) | 14.1%<br>(12.0-16.1%) | 7.9%<br>(7.0-8.7%) |
| Live in group home | 8.0%<br>(5.4-10.7%) | 4.1%<br>(3.0-5.3%) | 3.0%<br>(1.8-4.1%) | 7.0%<br>(5.5-8.5%) | 5.2%<br>(4.5-6.0%) |
| Other | 0.5%<br>(0.0-1.2%) | 4.7%<br>(3.5-5.9%) | 4.5%<br>(3.1-6.0%) | 4.2%<br>(3.0-5.3%) | 4.0%<br>(3.4-4.7%) |
| Diagnoses |  |  |  |  |  |
| Autism | 28.8%<br>(24.4-33.2%) | 15.4%<br>(13.3-17.5%) | 15.4%<br>(13.0-17.9%) | 22.3%<br>(19.9-24.7%) | 19.1%<br>(17.8-20.4%) |
| Down syndrome | 42.7%<br>(37.9-47.5%) | 29.2%<br>(26.6-31.8%) | 30.5%<br>(27.3-33.6%) | 21.3%<br>(18.9-23.7%) | 28.5%<br>(27.1-30.0%) |
| Fragile X | 7.3%<br>(4.8-9.8%) | 3.9%<br>(2.8-5.0%) | 1.9%<br>(1.0-2.8%) | 5.3%<br>(4.0-6.6%) | 4.3%<br>(3.6-4.9%) |
| Cerebral Palsy | 10.7%<br>(7.7-13.7%) | 8.3%<br>(6.7-9.9%) | 3.7%<br>(2.4-5.0%) | 5.5%<br>(4.2-6.8%) | 6.6%<br>(5.8-7.4%) |
| Other ID | 13.2%<br>(9.9-16.4%) | 44.9%<br>(42.1-47.8%) | 49.2%<br>(45.8-52.6%) | 47.6%<br>(44.7-50.5%) | 43.1%<br>(41.5-44.7%) |

<sup>a</sup>Respondents from 181 Special Olympics programs across 138 countries

Globally, 76.2% (74.8-77.6%) of participants received a COVID-19 vaccination while 49.5% (47.9-51.2%) received a COVID-19 booster (Table 2). Other ID (81.0%, 54.2%), Autism (76.1%, 46.6%) and Down syndrome (75.0%, 49.8%) were the diagnoses with higher levels of vaccine and booster receipt, respectively. COVID-19 vaccination status varied by economic levels with UMIC and HIC respondents having the highest rates (93.0% (91.2-94.7%) and 93.5% (92.1-95.0%), respectively), while LIC respondents reported the lowest proportion of vaccination (38.0% (33.3-42.7%)). This trend continued for COVID-19 booster vaccinations with UMIC and HIC respondents having the highest proportion (67.0% (63.8-70.2%) and 75.2% (72.7-77.7%), respectively) and LIC respondents with the lowest proportion (14.7% (11.3-18.1%)). Figure 1 displays the receipt of COVID-19 vaccinations and boosters by World Bank economic levels.

**Table 2.**
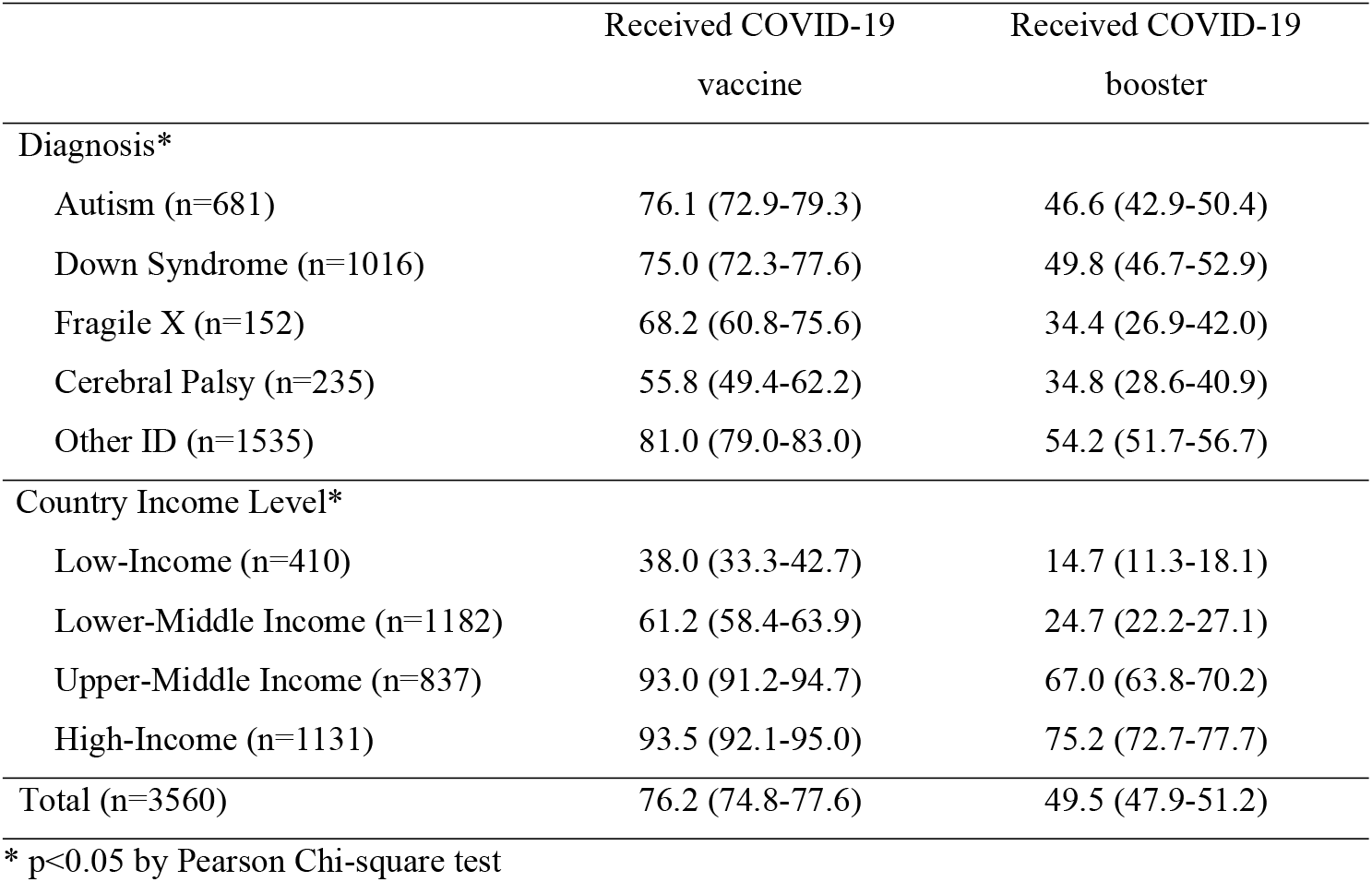
Receipt of COVID-19 Vaccine and Booster by Intellectual Disability and Country Income Level (%, 95% MoE)

**Fig 1.**
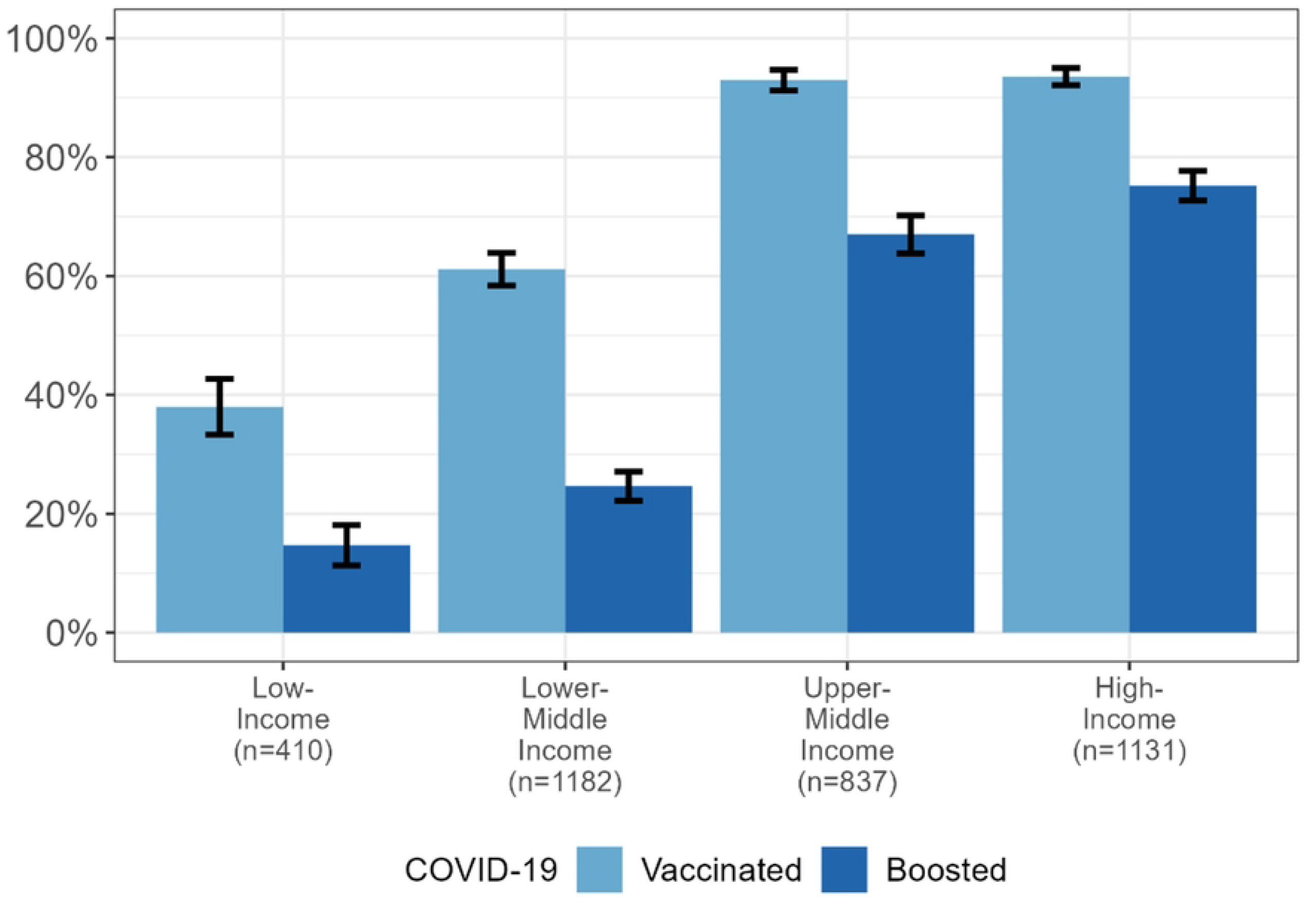
Receipt of COVID-19 Vaccine and Booster by Country Economic Income Level.

Univariate models of vaccination identified age and economic level to be positively associated with vaccination while living with family and having fragile X syndrome or cerebral palsy were inversely associated with vaccination (Table 3). In the multivariate logistic regression model, age (OR=1.04 per year [1.03, 1.05]), country economic level (OR=3.12 [2.81, 3.48]), and living with family (OR=0.70 [0.53, 0.92]) remained associated with vaccination status.

**Table 3.**
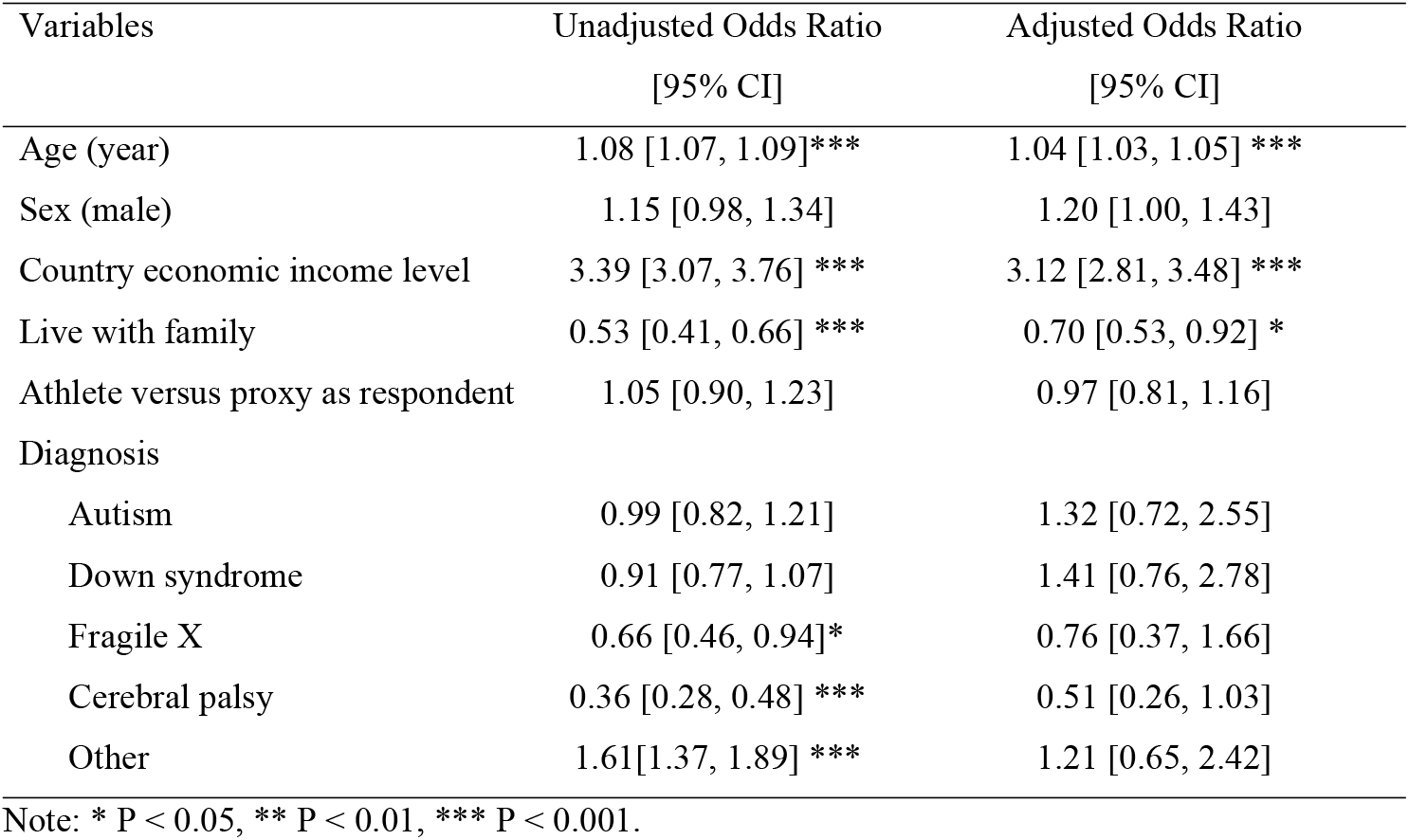
Univariate and multivariate logistic regression models of COVID-19 vaccination among Special Olympics global survey respondents (n=3560).

| Variables | Unadjusted Odds Ratio<br>[95% CI] | Adjusted Odds Ratio<br>[95% CI] |
| --- | --- | --- |
| Age (year) | 1.08 [1.07, 1.09]*** | 1.04 [1.03, 1.05] *** |
| Sex (male) | 1.15 [0.98, 1.34] | 1.20 [1.00, 1.43] |
| Country economic income level | 3.39 [3.07, 3.76] *** | 3.12 [2.81, 3.48] *** |
| Live with family | 0.53 [0.41, 0.66] *** | 0.70 [0.53, 0.92] * |
| Athlete versus proxy as respondent | 1.05 [0.90, 1.23] | 0.97 [0.81, 1.16] |
| Diagnosis |  |  |
| Autism | 0.99 [0.82, 1.21] | 1.32 [0.72, 2.55] |
| Down syndrome | 0.91 [0.77, 1.07] | 1.41 [0.76, 2.78] |
| Fragile X | 0.66 [0.46, 0.94]* | 0.76 [0.37, 1.66] |
| Cerebral palsy | 0.36 [0.28, 0.48] *** | 0.51 [0.26, 1.03] |
| Other | 1.61[1.37, 1.89] *** | 1.21 [0.65, 2.42] |

Note: \* P < 0.05, \*\* P < 0.01, \*\*\* P < 0.001.

Globally, among those not vaccinated for COVID-19, respondents in LLMICs reported a near threefold higher willingness to vaccinate (74.1% (70.9-77.3%)) compared to UMHICs (26.5% (19.0-34.0%)) (χ^2^ = 111.4, *p* < 0.001) (Table 4). The primary reasons to not vaccinate differed among adults with ID in LLMICs and UMHICs (p<0.05 by Pearson Chi-square test). In LIC, the most common reason for not vaccinating was having no access to vaccination (41.2% (29.5-52.9%)). Globally, the most common reasons for not vaccinating were concerns about side effects (42.3%, (36.5-48.1%)) and parent/guardian not wanting the adult with ID to vaccinate (31.5% (26.1-37.0%)).

**Table 4.** COVID-19 Vaccination Acceptance Among Unvaccinated Respondents and Reasons to Not Vaccinate (%, 95% Margin of Error).

|  | Low- Income | Lower-Middle Income | Upper-Middle Income | High-Income | Total |
| --- | --- | --- | --- | --- | --- |
| Plan to vaccinate | n=253 | n=458 | n=59 | n=73 | n=843 |
| Yes | 73.1%<br>(67.7-78.6%) | 74.7%<br>(70.7-78.7%) | 35.6%<br>(23.4-47.8%) | 19.2%<br>(10.1-28.2%) | 66.7%<br>(63.5-69.8%) |
| No | 7.9%<br>(4.6-11.2%) | 5.2%<br>(3.2-7.3%) | 30.5%<br>(18.8-42.3%) | 53.4%<br>(42.0-64.9%) | 12.0%<br>(9.8-14.2%) |
| Not sure | 19.0%<br>(14.1-23.8%) | 20.1%<br>(16.4-23.8%) | 33.9%<br>(21.8-46.0%) | 27.4%<br>(17.2-37.6%) | 21.4%<br>(18.6-24.1%) |
| Reasons to not vaccinate | n=68 | n=116 | n= 38 | n=57 | n=279 |
| Vaccine not safe* | 16.2%<br>(7.4-24.9%) | 25.0%<br>(17.1-32.9%) | 34.2%<br>(19.1-49.3%) | 47.4%<br>(34.4-60.3%) | 28.7%<br>(23.4-34.0%) |
| Concerned about side effects* | 19.1%<br>(9.8-28.5%) | 40.5%<br>(31.6-49.5%) | 60.5%<br>(45.0-76.1%) | 61.4%<br>(48.8-74.0%) | 42.3%<br>(36.5-48.1%) |
| Vaccine will not work* | 7.4%<br>(1.1-13.6%) | 9.5%<br>(4.2-14.8%) | 34.2%<br>(19.1-49.3%) | 33.3%<br>(21.1-45.6%) | 17.2%<br>(12.8-21.6%) |
| Distrust government* | 7.4%<br>(1.1-13.6%) | 5.2%<br>(1.1-9.2%) | 18.4%<br>(6.1-30.7%) | 28.1%<br>(16.4-39.7%) | 12.2%<br>(8.3-16.0%) |
| Distrust vaccine manufacture* | 4.4%<br>(0.0-9.3%) | 0.9%<br>(0.8-2.5%) | 18.4%<br>(6.1-30.7%) | 28.1%<br>(16.4-39.7%) | 9.7%<br>(6.2-13.1%) |
| Prior negative vaccine experience* | 1.5%<br>(0.0-4.3%) | 6.0%<br>(1.7-10.4%) | 15.8%<br>(4.2-27.4%) | 19.3%<br>(9.1-29.5%) | 9.0%<br>(5.6-12.3%) |
| Not enough information about the vaccine | 7.4%<br>(1.1-13.6%) | 31.9%<br>(23.4-40.4%) | 18.4%<br>(6.1-30.7%) | 26.3%<br>(14.9-37.7%) | 22.9%<br>(18.0-27.9%) |
| No access to vaccine* | 41.2%<br>(29.5-52.9%) | 7.8%<br>(2.9-12.6%) | 2.6%<br>(0.0-7.7%) | 0.0%<br>(0.0-0.0%) | 13.6%<br>(9.6-17.6%) |
| Religious objection* | 1.5%<br>(0.0-4.3%) | 2.6%<br>(0.0-5.5%) | 2.6%<br>(0.0-7.7%) | 26.3%<br>(14.9-37.7%) | 7.2%<br>(4.1-10.2%) |
| Do not have transportation | 16.2%<br>(7.4-24.9%) | 0.9%<br>(0.0-2.5%) | 2.6%<br>(0.0-7.7%) | 0.0%<br>(0.0-0.0%) | 4.7%<br>(2.2-7.1%) |
| Don't know where to get vaccinated | 7.4%<br>(1.1-13.6%) | 0.0%<br>(0.0-0.0%) | 0.0%<br>(0.0-0.0%) | 0.0%<br>(0.0-0.0%) | 1.8%<br>(0.2-3.3%) |
| Do not like needles | 17.6%<br>(8.6-26.7%) | 31.0%<br>(22.6-39.5%) | 15.8%<br>(4.2-27.4%) | 17.5%<br>(7.7-27.4%) | 22.9%<br>(18.0-27.9%) |
| Parent/ guardians do not want me to vaccinate | 26.5%<br>(16.0-37.0%) | 32.8%<br>(24.2-41.3%) | 23.7%<br>(10.2-37.2%) | 40.4%<br>(27.6-53.1%) | 31.5%<br>(26.1-37.0%) |
| Other | 1.5%<br>(0.0-4.3%) | 6.0%<br>(1.7-10.4%) | 7.9%<br>(0.0-16.5%) | 12.3%<br>(3.8-20.8%) | 6.5%<br>(3.6-9.3%) |

\* P < 0.05 by Pearson Chi-square test

## Discussion

This study identified an association between country economic level and COVID-19 vaccination and reasons for not accepting vaccine among adults with ID. Participants from LLMICs reported fewer COVID-19 vaccinations than those from UMHICs and a higher willingness to vaccinate among those who are not yet vaccinated. The most common reasons for not vaccinating offer opportunities to address concerns about vaccine side effects and family caregivers’ apprehension to vaccinate this high-risk population.

Over 75% of adults with ID in the current study were vaccinated against COVID-19 compared to 60% of the global population during the same time period (28). The relatively higher rates of COVID-19 vaccination among adults with ID in UMHICs suggest evidence of effective public health polices and favorable vaccine acceptance. At the time of this study, governments across the world implemented universal policies for vaccine delivery for vulnerable populations(29). The high rates of vaccination found in our study are compatible with favorable vaccine uptake among those with ID reported by Hatton et al. (83.5% vaccination rate) and Iadorala et al. (87.0% of the participants willing to get the COVID-19 vaccine) (30,31).

Despite overall high rates of vaccination among adults with ID, there were dramatic differences in vaccination across country economic levels. Lower overall vaccination in LICs compared to HICs is consistent with global COVID-19 vaccine dosages administered. As of January 2, 2022, *Our World in Data* reported higher levels of COVID-19 vaccine doses administered among UMICs (169.73 per 100 people) and HICs (166.18) compared to LMICs (84.12) and LICs (10.67)(32). Such findings suggest a lack of resources and infrastructure to vaccinate large populations among many LLMICs (33). As shown by the rates of under-vaccinated children in LLMICs, vaccine infrastructure may be inadequate for large scale pandemic response (33).

Among those not vaccinated, more adults with ID from LLMICs planned to receive the COVID-19 vaccine compared to UMHICs. Among the unvaccinated population overall, LLMICs generally show higher willingness to accept vaccines than higher income countries (34– 36). However, a recent study indicates growing pockets of vaccine hesitancy (37). As vaccines become more accessible to LLMIC, providers should focus communication on building trust, working with trusted stakeholders (e.g., community health workers), and engaging in broad community outreach to reduce vaccine hesitancy and rebuild confidence in vaccination programs for vulnerable and underserved communities (36).

Our findings document the most common reasons that adults with ID did not vaccinate across multiple levels of the sociological framework (38). Vaccination campaigns that target multiple levels of the socioecological model, focus on dialogue-based communication, and include community stakeholders yield better outcomes (37). At the individual level, concerns about vaccine side-effects and safety were top reasons for not receiving a COVID-19 vaccine across country economic levels. However, higher rates of respondents in UMHICs expressed concerns about side effects and safety than LLMICs. Those in UMHICs also expressed skepticism that the vaccine would work, which is consistent with prior research (39). At the interpersonal level of influence, the decision to vaccinate was significantly influenced by caregivers. Globally, parent/guardian restriction for vaccination was a primary reason those with ID were not vaccinating for COVID-19. These results complement Iadorola et al.’s (2022) study where those making decisions on behalf of a person with ID had more vaccine hesitancy than those with ID themselves (31). These collective findings highlight the need to provide vaccine outreach and education tailored to caregivers’ apprehension towards COVID-19 vaccination. Providing clear information about vaccine risks and incorporating trusted sources of information, including health care providers, may improve vaccine uptake worldwide for those with ID.

This study has several limitations. Findings are from a sample of adults with ID who participate in Special Olympics programming and may not be generalizable to the broader population of persons with ID. The study population includes 181 countries and states with Special Olympics affiliates but is not representative of all countries within economic levels. Because the survey was online, participants without internet access could not be included and likely contributed to underestimates of vaccination. As COVID-19 vaccination were self-reported, the vaccination type and dosage were not available for analysis. Nonetheless, this is the largest study of COVID-19 vaccination among people with ID to date that included representation from three-fourths of the world’s countries.

## Conclusion

This global study identified a significant association of country economic level on rates of COVID-19 vaccination and reasons for not accepting vaccine among adults with ID. Adults with ID from low and low-middle income countries reported fewer COVID-19 vaccinations, and indicated that the reason for not vaccinating was reduced access. Globally, COVID-19 vaccination levels among adults with ID were higher than the general population. Findings support the need to implement public health interventions that address family caregivers’ apprehension to vaccinate this high-risk population.

## Data Availability

A minimal data set is available upon request to the authors.

## Acknowledgments

The authors would like to thank the Special Olympics program staff around the world for their assistance in collecting this information.

